# Optimal control of the COVID-19 pandemic with non-pharmaceutical interventions

**DOI:** 10.1101/2020.04.22.20076018

**Authors:** T. Alex Perkins, Guido España

## Abstract

The COVID-19 pandemic has forced societies across the world to resort to social distancing to slow the spread of the SARS-CoV-2 virus. Due to the economic impacts of social distancing, there is growing desire to relax these measures. To characterize a range of possible strategies for control and to understand their consequences, we performed an optimal control analysis of a mathematical model of SARS-CoV-2 transmission. Given that the pandemic is already underway and controls have already been initiated, we calibrated our model to data from the US and focused our analysis on optimal controls from May 2020 through December 2021. We found that a major factor that differentiates strategies that prioritize lives saved versus reduced time under control is how quickly control is relaxed once social distancing restrictions expire in May 2020. Strategies that maintain control at a high level until summer 2020 allow for tapering of control thereafter and minimal deaths, whereas strategies that relax control in the short term lead to fewer options for control later and a higher likelihood of exceeding hospital capacity. Our results also highlight that the potential scope for controlling COVID-19 until a vaccine is available depends on epidemiological parameters about which there is still considerable uncertainty, including the basic reproduction number and the effectiveness of social distancing. In light of those uncertainties, our results do not constitute a quantitative forecast and instead provide a qualitative portrayal of possible outcomes from alternative approaches to control.

## 1 Introduction

The first known case of COVID-19 in the United States arrived from China on January 15, 2020 [22]. In the weeks that followed, deficiencies in testing allowed the virus that causes this disease to spread largely undetected [41], with the number of reported cases growing to 429,319 by April 8, 2020 [36]. Social distancing—such as closing schools, working from home, and sheltering in place—has been adopted widely and appears capable of impacting transmission [26,12]. These practices have tremendous economic and social consequences, however, which has precipitated growing desire to relax them [16].

A danger in relaxing the use of social distancing is that the virus could resurge in its absence [44]. A safe, effective, and widely used vaccine would obviate the need for social distancing, but there are many challenges associated with developing a vaccine against COVID-19 [30] and one is not likely to be available until at least spring 2021 [4]. Strategies for successfully controlling COVID-19 until then will depend on a suite of non-pharmaceutical interventions (NPIs) [3], including some degree of social distancing but also diagnostic testing, contact tracing, and case isolation [5].

In models of pathogen transmission based on mass-action assumptions, NPIs all act in a similar way by reducing the transmission coefficient. This parameter, often denoted *β*, reflects the product of the rate of contact between susceptible and infectious people and the probability of transmission upon contact [24]. Due to the extent to which they reduce contact, full-scale lockdowns appear to be the most effective strategy for reducing transmission available at present [18]. Until alternative NPI-based strategies can be implemented that are similarly effective but less disruptive economically, it is urgently important to determine the minimal extent to which contact must be reduced to achieve public health objectives.

Optimal control theory offers a way to understand how to apply one or more time-varying control measures to a nonlinear, dynamical system in such a way that a given objective is optimized [28]. These techniques have been widely applied to a variety of pathogen transmission systems before (e.g., [6,33,11,2]), including in the context of a pandemic of a respiratory pathogen (e.g., [29,48,46]). The latter studies indicate that the level of NPI-based control required is dependent on model parameters, and that application of NPIs can be required at a high level and for a long duration in the absence of a vaccine.

Here, we apply optimal control theory to determine optimal strategies for the implementation of NPIs to control COVID-19, with a focus on the US. An optimal strategy in this sense involves weighing the relative costs of control and COVID-related mortality, and finding an approach to control that minimizes that combined cost. Other optimal control analyses of COVID-19 are beginning to emerge [14,31,40,43,45], although these have been less focused on any particular geographic setting. One contextual feature of the US that our analysis considers is the level of control that was implemented early in the pandemic. We account for this, because our objective is less about understanding what level of control would have been optimal earlier on and more about understanding what level of control would be optimal going forward, conditional on what has happened already. In addition, we emphasize the sensitivity of our results to epidemiological parameters that are not yet well characterized but appear influential in determining what range of impacts of NPIs on COVID-19 are possible. All code and data used in our analysis is available at http://github.com/TAlexPerkins/covid19optimalcontrol.

## 2 Methods

### 2.1 Model

We modeled SARS-CoV-2 transmission according to a system of ordinary differential equations,

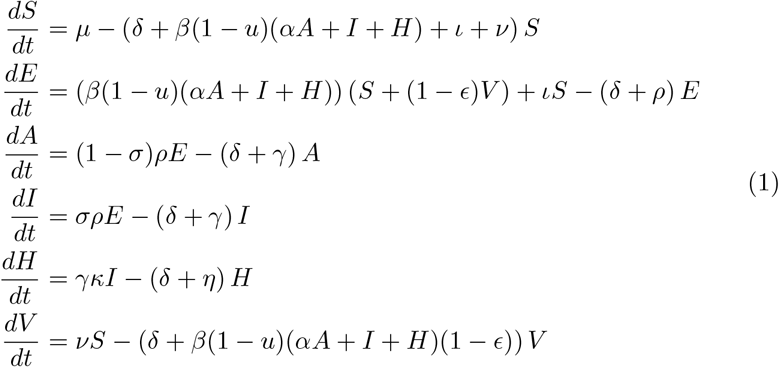

with variables and parameters defined in Tables 1 and 2.

**Table 1.** State variables in the model.

| Symbol | Definition |
| --- | --- |
| $S$ | Susceptible individuals |
| $E$ | Exposed but not yet infectious |
| $A$ | Asymptomatic infections |
| $I$ | Symptomatic infections |
| $H$ | Hospitalized infections |
| $V$ | Vaccinated individuals who have not been infected |
| $u$ | Control with non-pharmaceutical interventions |

**Table 2.** Model parameters. Parameterization of *β, δ, Δ*_−_, *Δ*_+_, *µ, ν, τ*_*ν*_, *H*_max_, and *u*_max_ is explained in further detail in Section 2.4. The timescale of all parameters related to time is daily.

| Symbol | Definition | Min | Mid | Max | Source |
| --- | --- | --- | --- | --- | --- |
| $\alpha$ | Relative infectiousness of asymptomatic infections | 0.046 | 0.602 | 0.981 | [41] |
| $\beta$ | Transmission coefficient | 0.514 | 0.600 | 0.686 | [39] |
| $\gamma$ | Rate of progression through infectious stage | 0.125 | 0.31 | 0.5 | [10] |
| $\delta$ | Baseline per capita death rate | - | $3.18 \times 10^{-5}$ | - | [32] |
| $\Delta_-$ | Minimum probability of death following hospitalization | 0.0865 | 0.104 | 0.1213 | [9] |
| $\Delta_+$ | Maximum probability of death following hospitalization | 0.233 | 0.28 | 0.328 | [38] |
| $\epsilon$ | Per exposure protection from vaccination | 0.5 | 0.8 | 0.9 | Assumed |
| $\eta$ | Rate of progression through hospitalization | 0.0485 | 0.075 | 0.172 | [1] |
| $\iota$ | Importation rate of infected people | $1.07 \times 10^{-7}$ | $3.24 \times 10^{-7}$ | $9.7 \times 10^{-7}$ | Calibrated |
| $\kappa$ | Probability of hospitalization among symptomatic infections | 0.207 | 0.26 | 0.314 | [9] |
| $\mu$ | Birth rate | - | $3.18 \times 10^{-5}$ | - | [32] |
| $\nu$ | Rate at which vaccines are administered | 0.001516 | 0.00197 | 0.002467 | [7,8] |
| $\rho$ | Rate of progression to infectiousness following infection | 0.166 | 0.2 | 0.333 | [10] |
| $\sigma$ | Proportion of infections that result in symptoms | 0.5 | 0.82 | 0.9 | [34] |
| $\tau_\nu$ | Day on which vaccination begins | 365 | 456 | 548 | [4] |
| $\omega$ | Proportion of deaths observed | 0.5 | - | 0.8 | Assumed |
| $h$ | Steepness of increase in death probability when $H_{\max}$ is exceeded | 1,403 | 701 | 350 | Assumed |
| $H_{\max}$ | Maximum hospital capacity | - | $2.74 \times 10^{-3}$ | - | [50] |
| $u_{\max}$ | Maximum value of the control | 0.5 | 0.7 | 0.9 | [23] |

In the model, all individuals are initially susceptible, *S*, with infections introduced at a constant rate, *ι*, from outside the population. Individuals then transition to the exposed class, *E*, where they reside for an average of *ρ*^−1^ days. A proportion *σ* experience a symptomatic infection, *I*. The remainder experience an asymptomatic infection, *A*, and have a fraction, *α*, of the infectiousness of symptomatic infections. Individuals reside in the *I* and *A* classes for an average of *γ*^−1^ days. All asymptomatic infections and a proportion 1 − *κ* of symptomatic infections then recover and become fully immune to subsequent infection. The remaining proportion *κ* of symptomatic infections transition to the hospitalized class, *H*, from which they exit through either recovery or death after an average of *η*^−1^ days.

Rather than track deaths due to COVID-19, *D*(*t*), as a state variable in eqn. (1), we assume that they follow directly from hospitalizations, *H*(*t*), according to

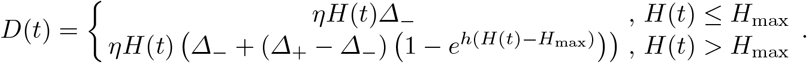

This results in the probability of death moving beyond a minimum of *Δ*_−_ towards a maximum of *Δ*_+_ as *H*(*t*) exceeds *H*_max_, as illustrated in Figure 1. The motivation for this choice is to account for the possibility that patients could experience increased mortality when the demand for certain resources, such as intensive care unit beds or ventilators, exceeds their availability. One other optimal control analysis of COVID-19 has incorporated a similar phenomenon [43], albeit with a different functional form.

**Fig. 1.**
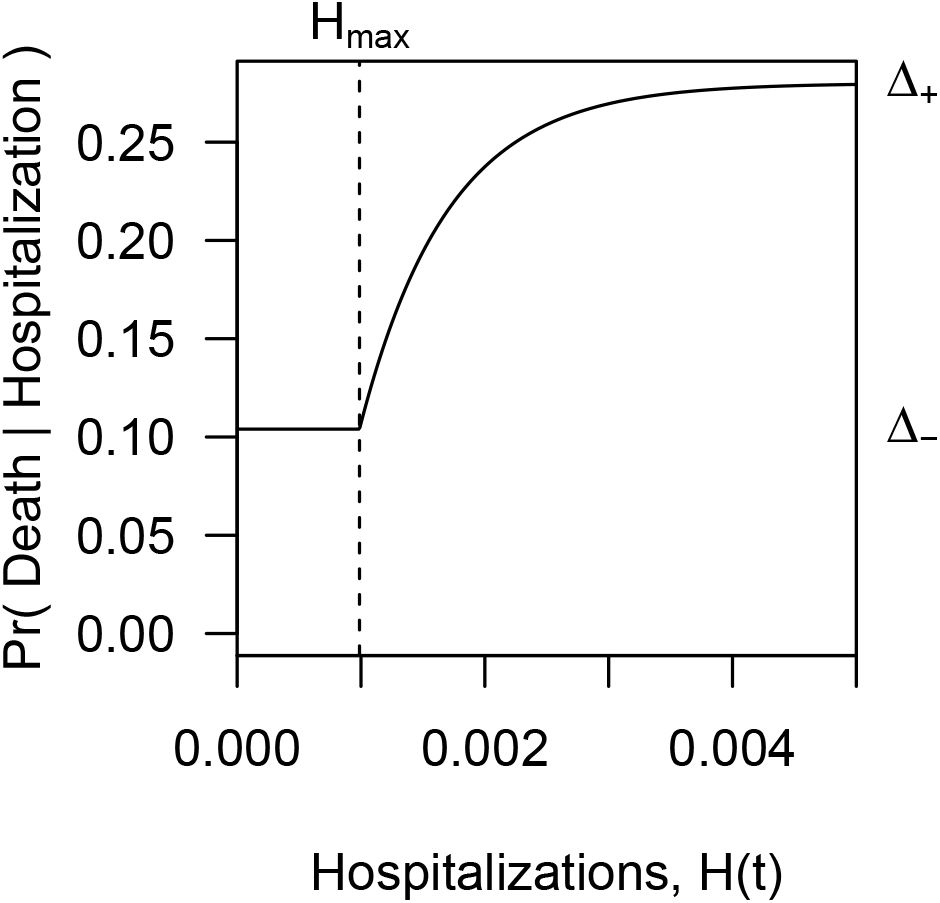
Relationship between the number of hospitalizations and the probability of death from COVID-19 among hospitalized patients. The parameters *Δ*_−_ and *Δ*_+_ represent lower and upper bounds on the probability of death, and *H*_max_ represents the hospital capacity above which the probability of death exceeds *Δ*_−_. Hospitalizations are quantified as a proportion of the overall population.

Individuals who are recovered and immune are not followed explicitly, as the model’s assumption of density-dependent transmission only requires specification of susceptible and infectious classes in the transmission term. Due to our assumption that rates of birth, *µ*, and death due to reasons other than COVID-19, *δ*, are equal, changes in population size over the course of the epidemic are modest, making the distinction between density- and frequency-dependent transmission negligible.

The primary form of control in the model is achieved through the variable *u*, which represents a proportional reduction in the transmission coefficient, *β*. As is standard for mass-action models of directly-transmitted pathogens, *β* reflects the product of the rate at which susceptible and infectious individuals come into contact and the probability of transmission given that a contact has occurred [24]. Thus, a wide range of NPIs could result in changes in *u*, including school closures, work from home policies, and shelter in place mandates, as well as more targeted approaches, such as isolation based on self-awareness of symptoms or contact tracing. In addition to control through *u*, the *V* class represents individuals who have been vaccinated, with individuals entering *V* from *S* at rate *ν* beginning on day *τ*_*ν*_. We assume that vaccination may not provide complete protection, resulting in vaccinated individuals becoming infected at a fraction 1 − *ϵ* of the rate at which fully susceptible individuals become infected.

### 2.2 Basic reproduction number, *R*_0_

At its core, the behavior of this model is similar to that of an SEIR model with demography. Because analyses of the transient and asymptotic properties of this class of models are plentiful in textbooks and elsewhere (e.g., [24]), we omit such an analysis here. We do, however, derive a formula to express the basic reproduction number, *R*_0_, as a function of model parameters. This relationship plays a role in how we parameterize the model.

We use the next-generation method [15] to obtain a formula describing *R*_0_ as a function of model parameters. This method depends on matrices ℱ and 𝒱, whose elements are defined as the rates at which secondary infections increase the *i*^th^ compartment and the rates at which disease progression, death, and recovery decrease the *i*^th^ compartment, respectively. For “disease compartments” *E, A, I*, and *H*, these matrices are defined as

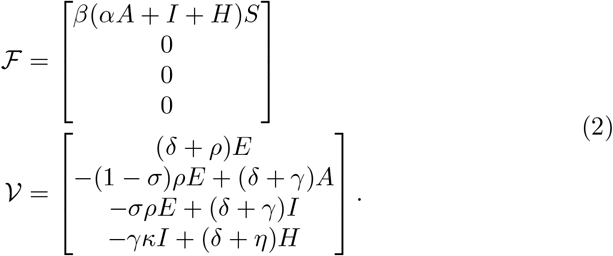

These matrices are then used to define two others,

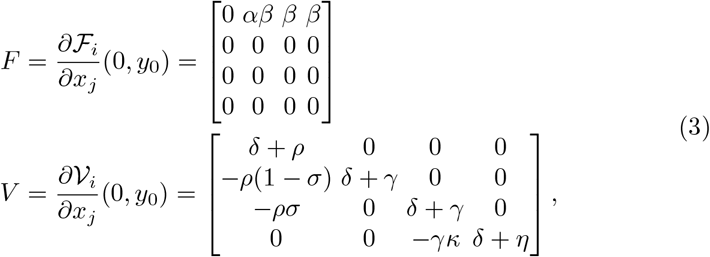

where *x* represents disease compartments and *y* non-disease compartments.

The inverse of *V* is

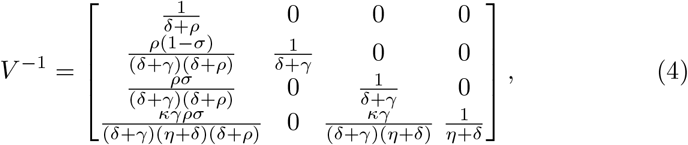

which, along with *F*, defines the matrix *K* = *FV* ^−1^. Specifying

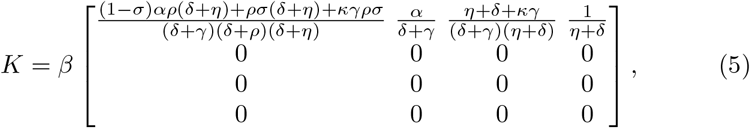

we obtain

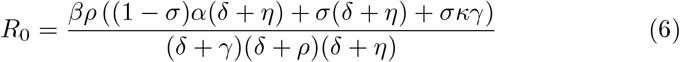

as the maximum eigenvalue of *K*.

### 2.3 Optimal control problem

The optimal control problem is to minimize the objective functional

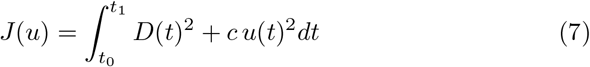

subject to the constraints of the state dynamics described in eqn. (1) and the initial conditions that *S*(*t*_0_) = 1 and all other state variables equal zero at time *t*_0_. The parameter *c* weights the extent to which the control, *u*(*t*), is prioritized for minimization relative to deaths, *D*(*t*). Squared terms for *D*(*t*) and *u*(*t*) are chosen both for mathematical convenience in the case of *u*(*t*) and to more heavily penalize solutions for *u*(*t*) that permit relatively high values of *D*(*t*) or *u*(*t*) at any given time.

To find the optimal control, *u*^*∗*^(*t*), that minimizes *J*(*u*), we follow standard results from optimal control theory applied to systems of ordinary differential equations [28]. These techniques make use of Pontryagin’s Maximum Principle to determine the pointwise minimum of the Hamiltonian of the system, ℋ, using adjoint variables, *λ*, that correspond to each of the state variables.

To solve for the adjoint variables, we first define the Hamiltonian,

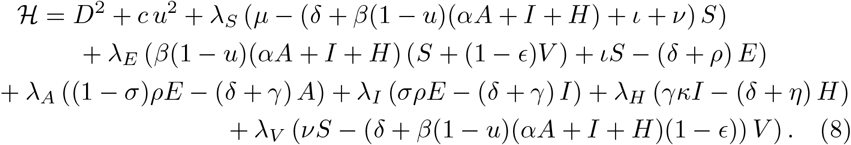

We then define differential equations describing the behavior of each adjoint variable as the negative of the partial derivative of ℋ with respect to the state variable corresponding to each adjoint variable. This yields

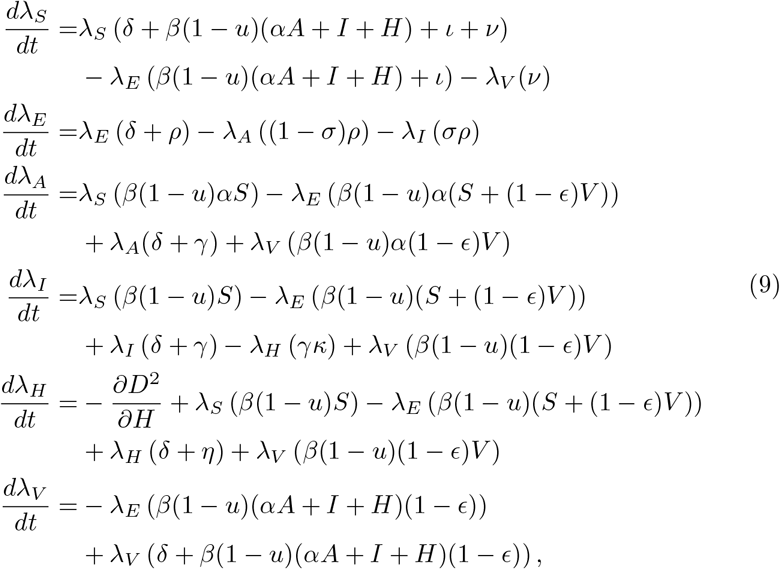

where

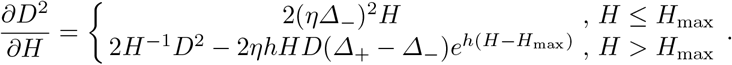

Eqn. (9) can be solved backwards in time with transversality conditions at time *t*_1_ equal to zero for each adjoint variable.

To find the pointwise optimal control, *u*^*∗*^, we find the value of *u* that minimizes 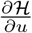, which yields

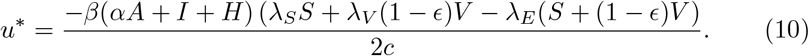

The optimal control is subject to a lower bound of 0 and an upper bound of *u*_max_, with those values used for *u*^*∗*^ whenever the right hand side of eq. (10) yields values outside those bounds.

To find *u*^*∗*^(*t*) numerically, we use the forward-backward sweep method [28], which involves first solving for the state variables forward in time, next solving for the adjoint variables backward in time, and then plugging the solutions for the relevant state and adjoint variables into eqn. (10), subject to bounds on *u*(*t*). As is often the case for optimal control problems [28], we found that we needed to perform these steps iteratively and with a convex combination of controls across iterations to achieve convergence. Specifically, we repeated the forward-backward sweep process 50 times, after which we took newly proposed solutions of *u*^*∗*^(*t*) as the average of the 20 most recent solutions, until the algorithm was stopped after 2,000 iterations. We assessed convergence with a statistic defined as

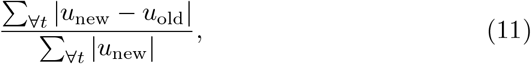

where smaller values indicate better convergence. All numerical solutions were obtained with a Runge-Kutta 4 routine implemented with the ode function from the deSolve package [47] in R.

### 2.4 Model parameterization

In Table 2, we specify low, intermediate, and high values of each parameter. For some parameters, estimates matching our parameter definitions were taken from other studies. For other parameters, additional steps were necessary to match our parameter definitions. We elaborate on the latter below.

#### Transmission coefficient, β

We based values of this parameter on assumptions about *R*_0_ by solving for *β* as a function of *R*_0_ and other parameters in eqn. (6). Because estimates of *R*_0_ for COVID-19 vary widely [39], we chose values that span a range of estimates that may be applicable to the US.

#### Background birth and death rates, µ and δ

We parameterized *µ* consistent with a rate of 3,791,712 births in a population of 331 million in the US in 2018 [32]. To achieve a constant population size in the absence of COVID-19, we set *δ* equal to *µ*.

#### Probability of death among hospitalized cases, Δ and h

We assume that *Δ*_−_ is equal to early estimates from the US (2.6%) [9] and that *Δ*_+_ is equal to estimates from Italy (7.2%) [38]. Estimates of how quickly *Δ*_+_ might be approached have not been made empirically, so the value of *h* was assumed. The intermediate value of *h* = 701 corresponds to an increase of *H* of 50% beyond *H*_max_ resulting in 50% of the maximum increase from *Δ*_−_ to *Δ*_+_.

#### Progression through hospitalization, η

We used line-list data [1] to estimate a mean (13.2 days) and standard deviation (7.39 days) of the time between hospital admission and either discharge or death.

#### Timing of vaccine introduction, τ_ν_

Experts have stated that a vaccine against COVID-19 could be available to the public by spring 2021 [4]. We used April 1, 2021 as our default value for this parameter.

#### Vaccination rate, ν

In the 2009 H1N1 pandemic, 100 million doses of vaccine were administered between October 2009 and April 2010 [7,8]. Consistent with that, our default value of *ν* resulted in 0.197% of the population being vaccinated each day.

#### Hospital capacity, H_max_

We based our definition of this parameter on hospital beds and did not include other aspects of hospital resources, such as ICU beds, ventilators, or hospital staffing. Specifically, we adopted an estimate of 312,090 beds available for COVID-19 patients in the US by the Institute for Health Metrics and Evaluation [50].

#### Maximum effect of control, u_max_

A model incorporating survey data and age-based contact patterns estimated that social distancing could reduce transmission of SARS-CoV-2 by 73% [23]. Based on this, we used 0.7 as an intermediate value of this parameter and 0.5 and 0.9 as lower and upper values.

### 2.5 Model calibration

To obtain realistic behavior of the model, we calibrated it to match the cumulative number of reported deaths in the US within the first 100 days of 2020 (i.e., by April 9), which we obtained from the New York Times [36]. We focused our calibration on the parameter *ι*, due to the difficulty of empirically estimating the rate at which imported infections appear. To obtain a value of *ι* that resulted in the model matching the reported number of deaths, we simulated the model across 300 values of *ι* evenly spaced between 10^−12^ and 10^−4^ on a log scale, performed a linear interpolation of the simulated number of deaths across those values of *ι*, and found the value of *ι* that most closely matched reported deaths.

We calibrated the model under a total of 18 different parameter scenarios, crossing low, intermediate, and high values of *R*_0_ with low, intermediate, and high values of *u*_max_ and low and high values of *ω*. The latter represents the proportion of all deaths caused by COVID-19 that were reported. Because NPIs began going into effect in the US within the timeframe of this calibration period, we calibrated the model subject to an assumed pattern of *u*(*t*) through the first 100 days of 2020. We chose a logistic functional form for this, with a minimum of 0 and a maximum of *u*_max_. Two parameters that control the midpoint and slope of the increase from 0 to *u*_max_ were selected by an informal process of trial and error, with the goal of having the model’s predictions of deaths over time match the timing of reported deaths under all 18 parameter scenarios. We also used this process to select the date on which importations were initiated through *ι*.

## 3 Results

### 3.1 Model calibration

Under each of the 18 different scenarios we considered about *R*_0_, *u*_max_, and *ω*, our model was able to reproduce the numbers and timing of reported deaths in the US reasonably well (Figure 2). Different values of *ι* were required to do so under different values of those other three parameters, but all values of *ι* ranged 10^−7^ - 10^−6^ (Table 3). Taking into account the population of the US, this equates to a range of 33 - 331 imported infections per day. Higher values of *R*_0_ resulted in lower values of *ι*, given that fewer importations were required to generate the reported numbers of deaths when there was more local transmission. For similar reasons, *ι* was lower when *u*_max_ was lower. The value of *ω* had a negligible effect on *ι*.

**Table 3.** Calibrated estimates of the importation rate, *ι*, under 18 scenarios about the values of *R*_0_, *u*_max_, and *ω*.

| $R_0$ | $u_{\max}$ | $\omega$ | $\iota$ |
| --- | --- | --- | --- |
| 3 | 0.5 | 0.5 | $9.35 \times 10^{-7}$ |
| 3.5 | 0.5 | 0.5 | $3.16 \times 10^{-7}$ |
| 4 | 0.5 | 0.5 | $1.07 \times 10^{-7}$ |
| 3 | 0.7 | 0.5 | $9.69 \times 10^{-7}$ |
| 3.5 | 0.7 | 0.5 | $3.32 \times 10^{-7}$ |
| 4 | 0.7 | 0.5 | $1.14 \times 10^{-7}$ |
| 3 | 0.9 | 0.5 | $9.35 \times 10^{-7}$ |
| 3.5 | 0.9 | 0.5 | $3.16 \times 10^{-7}$ |
| 4 | 0.9 | 0.5 | $1.07 \times 10^{-7}$ |
| 3 | 0.5 | 0.8 | $9.69 \times 10^{-7}$ |
| 3.5 | 0.5 | 0.8 | $3.32 \times 10^{-7}$ |
| 4 | 0.5 | 0.8 | $1.14 \times 10^{-7}$ |
| 3 | 0.7 | 0.8 | $9.35 \times 10^{-7}$ |
| 3.5 | 0.7 | 0.8 | $3.16 \times 10^{-7}$ |
| 4 | 0.7 | 0.8 | $1.07 \times 10^{-7}$ |
| 3 | 0.9 | 0.8 | $9.69 \times 10^{-7}$ |
| 3.5 | 0.9 | 0.8 | $3.32 \times 10^{-7}$ |
| 4 | 0.9 | 0.8 | $1.13 \times 10^{-7}$ |

**Fig. 2.**
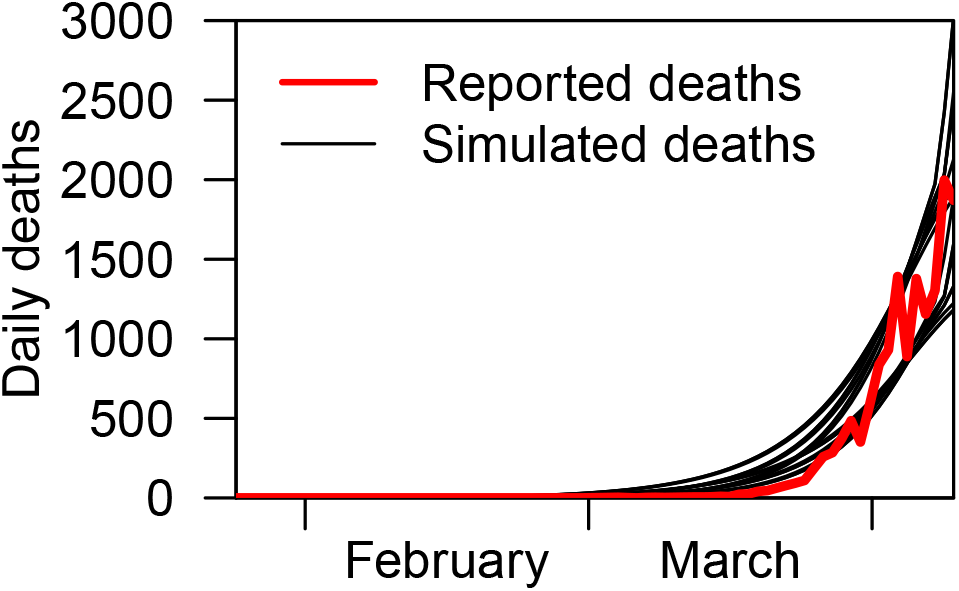
Reported (red) and simulated (black) numbers of daily deaths in the US resulting from model calibration under 18 different parameter scenarios (black lines).

In addition to *ι*, we also calibrated the date on which importations began (February 1) and *u*(*t*) during the first 100 days of 2020. Even though the first imported cases in the US began appearing before February 1 [22], it is not surprising that our model better matched the data with a slightly later start date, given that the model did not allow for increases in importation over time, as likely occurred [41]. The calibration of *u*(*t*) was consistent with *u*(*t*) starting at zero, reaching half of *u*_max_ on April 4, 2020, and increasing by a maximum of 0.075 *u*_max_ per day around that time. The timing of these changes is approximately two weeks later than changes in Google mobility data from the US [20], which may be a consequence of our model moving some individuals from exposure to death too quickly given that residence time in each compartment is exponentially distributed.

In general, we do not interpret any of the calibrated parameter values as reliable estimates of empirical quantities. Such an interpretation would require analyses that more carefully account for data-generating processes and sources of uncertainty. Rather, our objective in this calibration exercise was to ensure that the model’s behavior is reasonably consistent with empirical data, which we feel was accomplished.

### 3.2 Convergence

For our main results, we obtained solutions to *u*^*∗*^(*t*) under a total of 126 parameter combinations, crossing the 18 parameter sets in Table 3 with seven values of *c* spanning 10^−12^ to 10^−6^ by factors of ten. In one representative example (Figure 3, left), we observed that solutions of *u*^*∗*^(*t*) converged to a set of similar solutions after several hundred iterations. As expected, the objective functional decreased during those initial iterations and remained low thereafter (Figure 3, right). To assess convergence by the statistic in eqn. (11), we selected the iterations with the ten lowest values of *J* (*u*) in the last 100 iterations. Of 126 parameter combinations, 78% had final solutions of *u*^*∗*^(*t*) with convergence statistics below 10^−5^, 94% below 10^−2^, and all below 10^−1^.

**Fig. 3.**
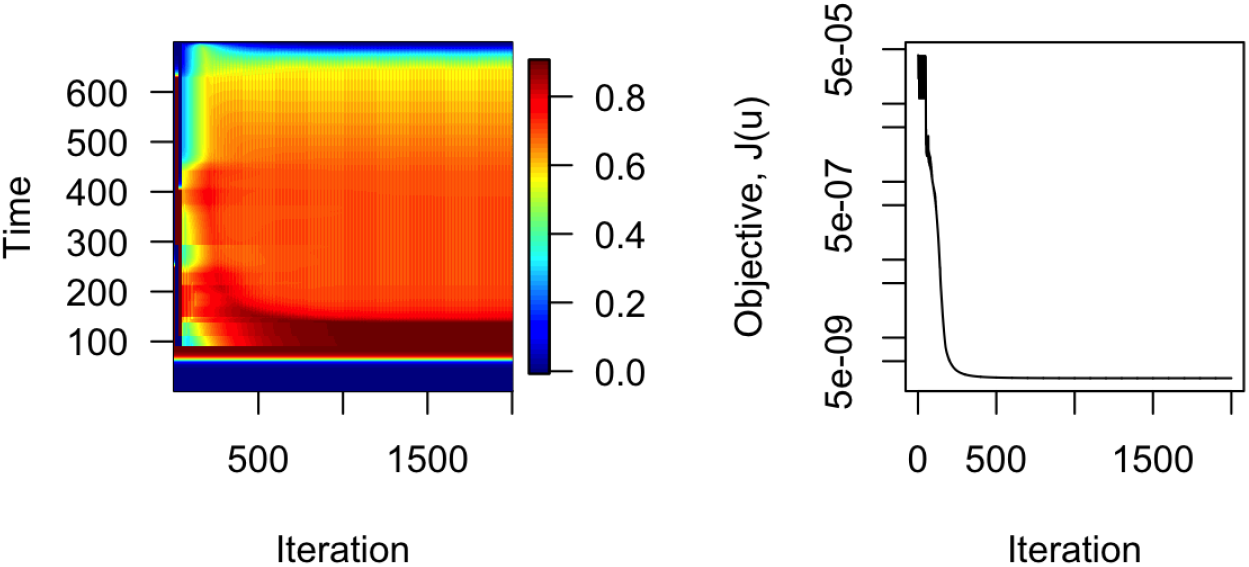
Convergence of solutions of *u*^*∗*^(*t*) under parameters *R*_0_ = 3, *u*_max_ = 0.9, *ω* = 0.8, and *c* = 10^−12^. Left: Colors indicate values of *u*^*∗*^(*t*) for each day in 2020 and 2021 across 2,000 iterations of the forward-backward sweep algorithm. Right: Across iterations, the value of the objective functional, *J* (*u*), decreased steadily until cycling for the remaining iterations.

### 3.3 Minimized objective functional under different parameters

Comparing values of the two components of the objective functional, 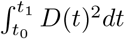 and 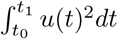, across different parameter values provides insight into the range of behavior of the model and its response to control. Because they are similar to the components of the objective functional but more easily interpretable, we describe effects of model parameters on 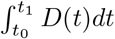 (cumulative deaths) and 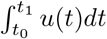 (cumulative time under control). The parameter *c*, which controls the weight of the two terms in *J* (*u*), led to a transition from minimizing cumulative deaths to minimizing cumulative time spent under control as it varied from 10^−9^ to 10^−6^ (Figure 4). The parameters *R*_0_ and *u*_max_ both influenced the extent to which cumulative deaths could be minimized. With a high value of *u*_max_, deaths could be kept relatively low across all values of *R*_0_ explored, provided that *c* ≤ 10^−10^. With a low value of *u*_max_ and a high value of *R*_0_, cumulative deaths could only be reduced by around 10% as *c* varied across its entire range from 10^−12^ to 10^−6^. The parameter *ω* had essentially no influence on the components of the objective functional (Figure 4).

**Fig. 4.**
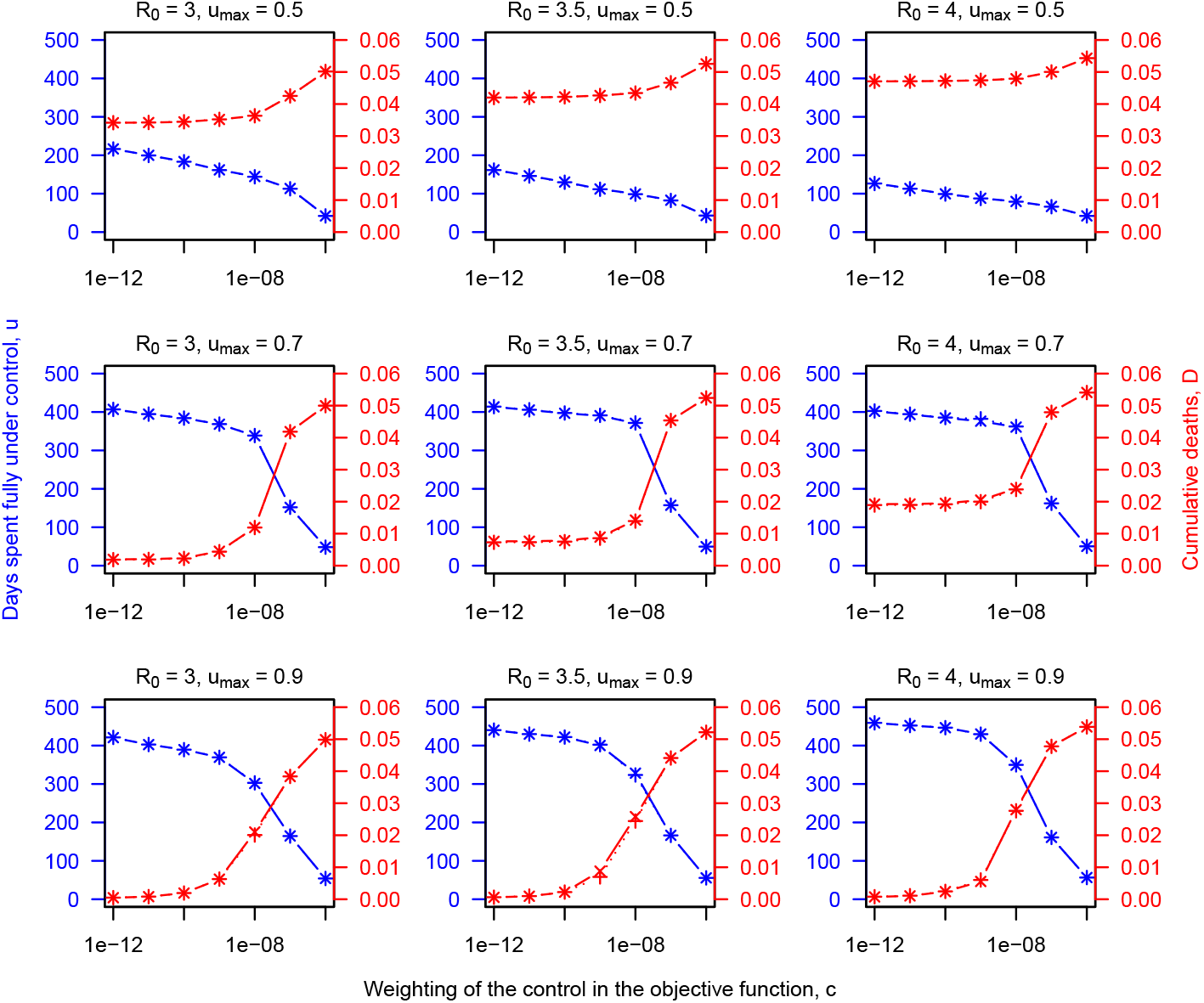
Dependence of time under control (blue) and cumulative deaths (red) on *c* (x-axis), *R*_0_ (columns), *u*_max_ (rows), and *ω* (markers). Deaths are quantified as a proportion of the overall population.

### 3.4 Optimal control over time

We first consider the scenario where *R*_0_ = 3 and *u*_max_ = 0.9, because those are the conditions under which control has the greatest potential to influence the pandemic. Under *c* = 10^−12^, *u*^*∗*^(*t*) remains at *u*_max_ until late June 2020, after which it settles down to around 75% of *u*_max_ until late 2021 (Figure 5). This results in hospitalizations dropping from their peak in April 2020 and remaining very low through 2021. The susceptible population remains very high and only begins eroding once a vaccine is introduced.

**Fig. 5.**
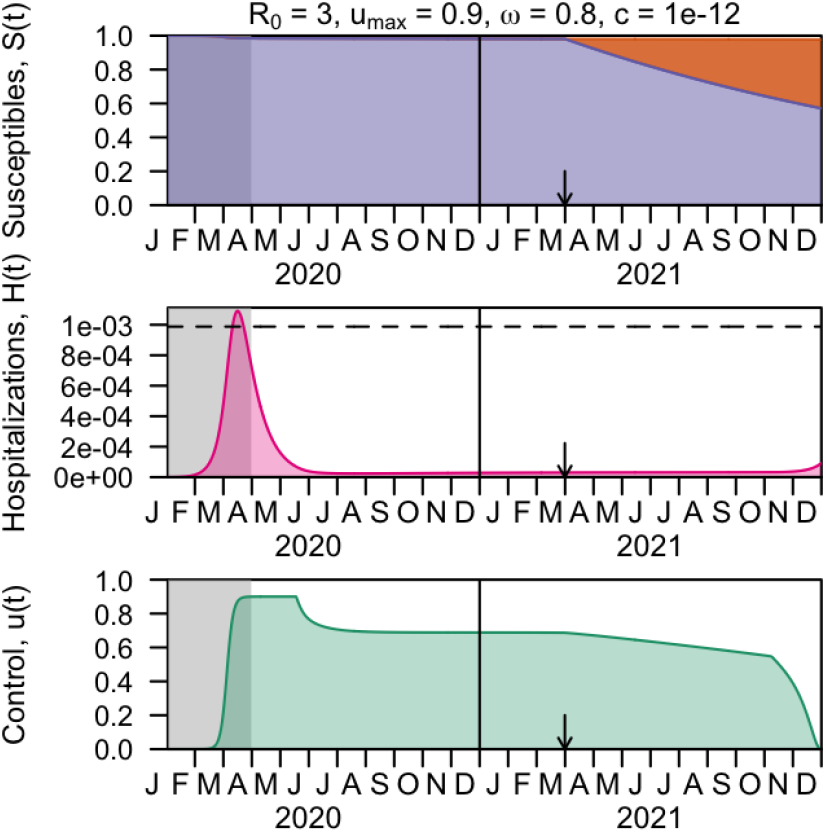
Optimal control under parameters with maximal ability to control the pandemic, with maximal weighting on minimization of deaths. Panels show the optimal control (bottom) and its impacts on the dynamics of hospitalized (middle) and susceptible (top) compartments with *R*_0_ = 3, *u*_max_ = 0.9, *ω* = 0.8, and *c* = 10^−12^. Vaccination is introduced at the time indicated by the arrow, with the vaccinated population (orange) reducing the susceptible population. Through April 2020 (gray shading), the value of the control is fixed according to its calibrated trajectory. The dashed horizontal line indicates hospital capacity.

With a higher value of *c* = 10^−9^, *u*^*∗*^(*t*) drops to around 50% of *u*_max_ in May 2020 (Figure 5). As a result, hospitalizations rebound and exceed hospital capacity by around a third in June and July before falling again, after *u*^*∗*^(*t*) returns to *u*_max_. From August 2020 onward, a steady decline in *u*^*∗*^(*t*) allows hospitalizations to be maintained at moderate levels before rebounding and exceeding hospital capacity once again for several months in 2021.

With the highest value of *c* = 10^−6^, *u*^*∗*^(*t*) drops almost to zero at the beginning of May 2020 (Figure. 7). This results in a rapid increase in hospitalizations, which is followed by an increase in *u*^*∗*^(*t*). By the end of June, the susceptible population has been depleted to the point that herd immunity begins to obviate the need for control and *u*^*∗*^(*t*) declines to zero by October 2020. During this large second wave in summer 2020, hospital capacity is ex-ceeded by more than 20-fold. This results in cumulative deaths equaling 5% of the population, which is approximately one order of magnitude greater than when *c* = 10^−9^ and two orders of magnitude greater than when *c* = 10^−12^.

**Fig. 6.**
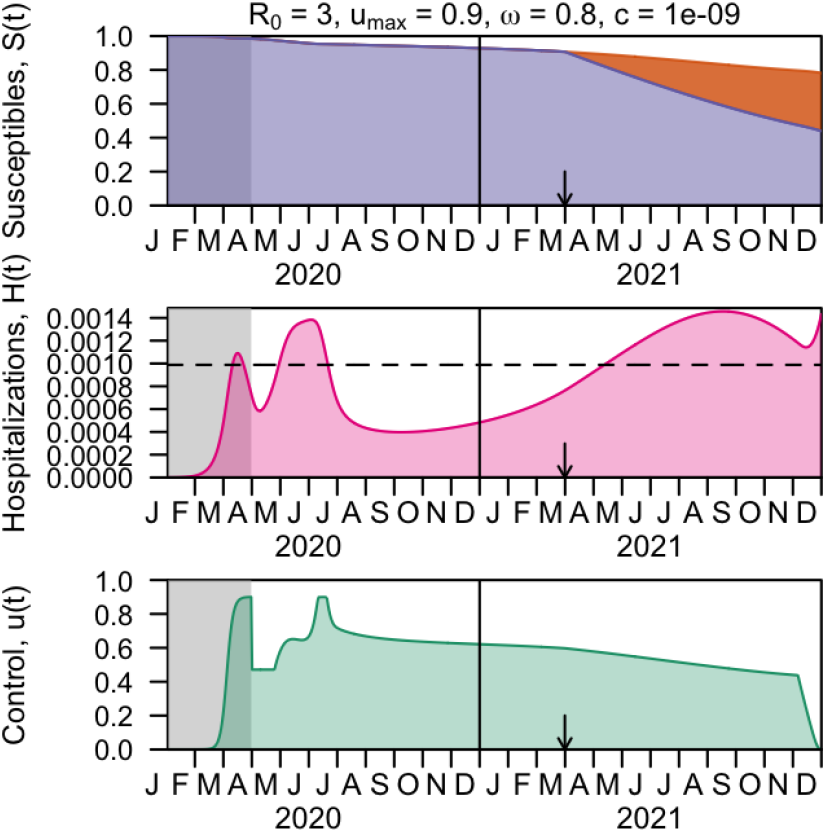
Optimal control under parameters with maximal ability to control the pandemic, with intermediate weighting on minimization of deaths. Panels show the optimal control (bottom) and its impacts on the dynamics of hospitalized (middle) and susceptible (top) compartments with *R*_0_ = 3, *u*_max_ = 0.9, *ω* = 0.8, and *c* = 10^−9^. Vaccination is introduced at the time indicated by the arrow, with the vaccinated population (orange) reducing the susceptible population. Through April 2020 (gray shading), the value of the control is fixed according to its calibrated trajectory. The dashed horizontal line indicates hospital capacity.

**Fig. 7.**
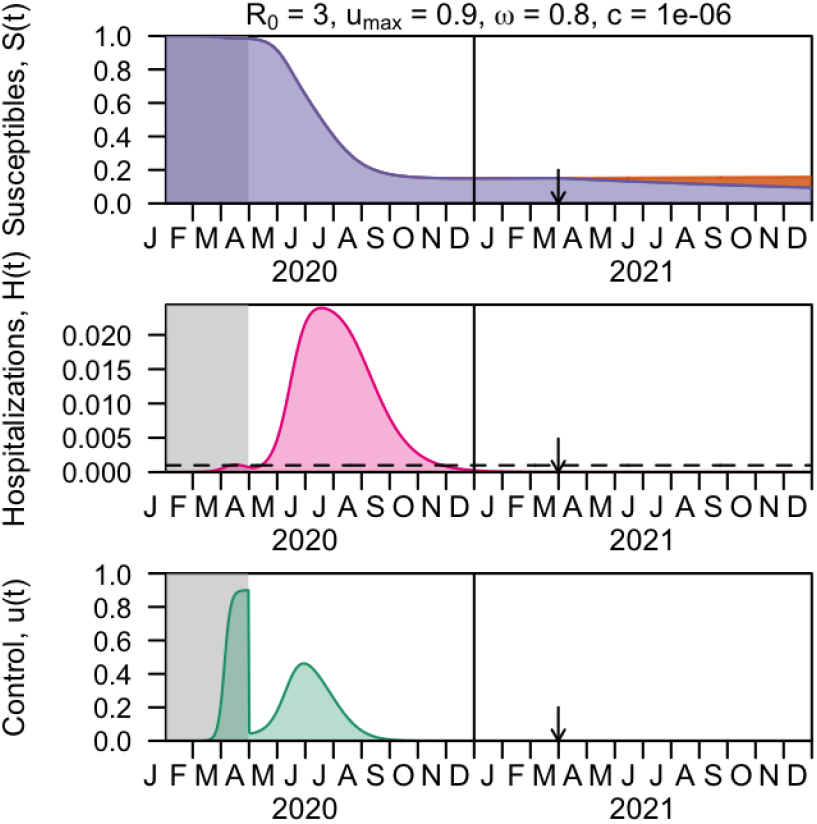
Optimal control under parameters with maximal ability to control the pandemic, with minimal weighting on minimization of deaths. Panels show the optimal control (bottom) and its impacts on the dynamics of hospitalized (middle) and susceptible (top) compartments with *R*_0_ = 3, *u*_max_ = 0.9, *ω* = 0.8, and *c* = 10^−9^. Vaccination is introduced at the time indicated by the arrow, with the vaccinated population (orange) reducing the susceptible population. Through April 2020 (gray shading), the value of the control is fixed according to its calibrated trajectory. The dashed horizontal line indicates hospital capacity.

Under other conditions about the transmissibility of the virus and the potential for NPIs to reduce transmission, control is less capable of curtailing the epidemic. With *R*_0_ = 3.5 and *u*_max_ = 0.7, hospitalizations peak in early summer 2020 at twice hospital capacity and take until December 2020 to fall below hospital capacity, even with the most aggressive value of *c* that we considered (10^−12^) (Figure 8). With *R*_0_ = 4, *u*_max_ = 0.5, and *c* = 10^−12^, hospitalizations peak in summer 2020 at thirty times hospital capacity (Figure 9). This results in extensive herd immunity and the relaxation of control in 2021, albeit at the cost of extensive deaths (Figure 4). Optimal controls under a wider variety of parameter combinations can be explored interactively at http://covid19optimalcontrol.crc.nd.edu.

**Fig. 8.**
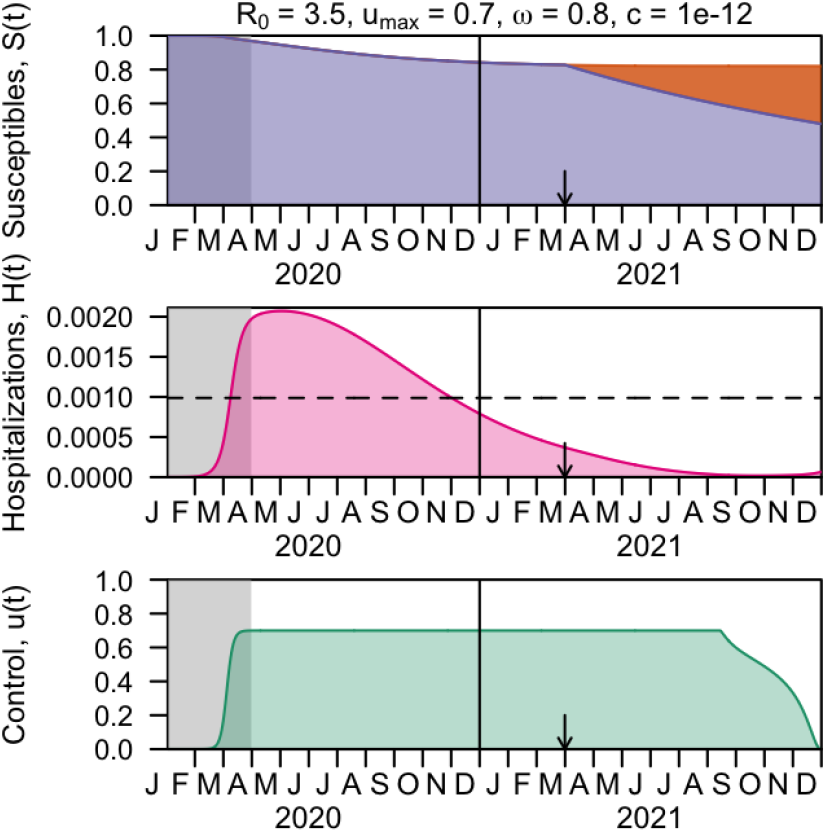
Optimal control under parameters with maximal ability to control the pandemic, with maximal weighting on minimization of deaths. Panels show the optimal control (bottom) and its impacts on the dynamics of hospitalized (middle) and susceptible (top) compartments with *R*_0_ = 3.5, *u*_max_ = 0.7, *ω* = 0.8, and *c* = 10^−12^. Vaccination is introduced at the time indicated by the arrow, with the vaccinated population (orange) reducing the susceptible population. Through April 2020 (gray shading), the value of the control is fixed according to its calibrated trajectory. The dashed horizontal line indicates hospital capacity.

**Fig. 9.**
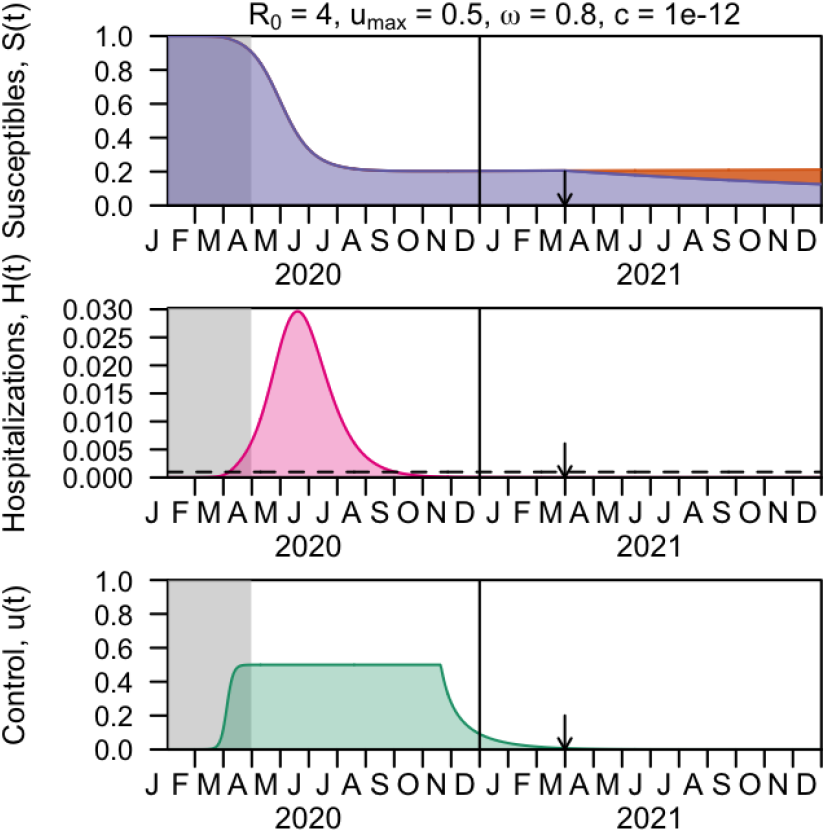
Optimal control under parameters with maximal ability to control the pandemic, with maximal weighting on minimization of deaths. Panels show the optimal control (bottom) and its impacts on the dynamics of hospitalized (middle) and susceptible (top) compartments with *R*_0_ = 4, *u*_max_ = 0.5, *ω* = 0.8, and *c* = 10^−12^. Vaccination is introduced at the time indicated by the arrow, with the vaccinated population (orange) reducing the susceptible population. Through April 2020 (gray shading), the value of the control is fixed according to its calibrated trajectory. The dashed horizontal line indicates hospital capacity.

### 3.5 Optimal control following different starting conditions

Our calibration procedure resulted in a single assumption about the trajectory of *u*(*t*) prior to April 30, 2020. Control during this period was fixed in analyses in Section 3.4, with the flexibility to define *u*^*∗*^(*t*) only allowed during the period from May 1, 2020 to December 31, 2021. Here, we explore how NPIs initiated one to three weeks earlier or later in spring 2020 would affect optimal controls in the period after. We focus this analysis on scenarios in which *c* = 10^−12^, which results in the optimal control problem seeking to minimize deaths as aggressively as any scenario that we explored. Consequently, we interpret changes in the optimal control in this section to reflect changes in constraints on what solutions of *u*^*∗*^(*t*) are possible, rather than changes in the balance between *D*(*t*)^2^ and *u*(*t*)^2^ in the optimization.

Across all combinations of *R*_0_, *u*_max_, and *ω* that we considered, cumulative deaths through 2020 and 2021 decrease when control begins earlier and increase when control begins later (Figure 10, red). In the scenario in which a delay in the initiation of control has the smallest effect (*R*_0_ = 3, *u*_max_ = 0.5), cumulative deaths increase by 10% with a three-week delay. In the scenario in which a delay in the initiation of control has the largest effect (*R*_0_ = 4, *u*_max_ = 0.9), cumulative deaths increase 28-fold with a three-week delay.

**Fig. 10.**
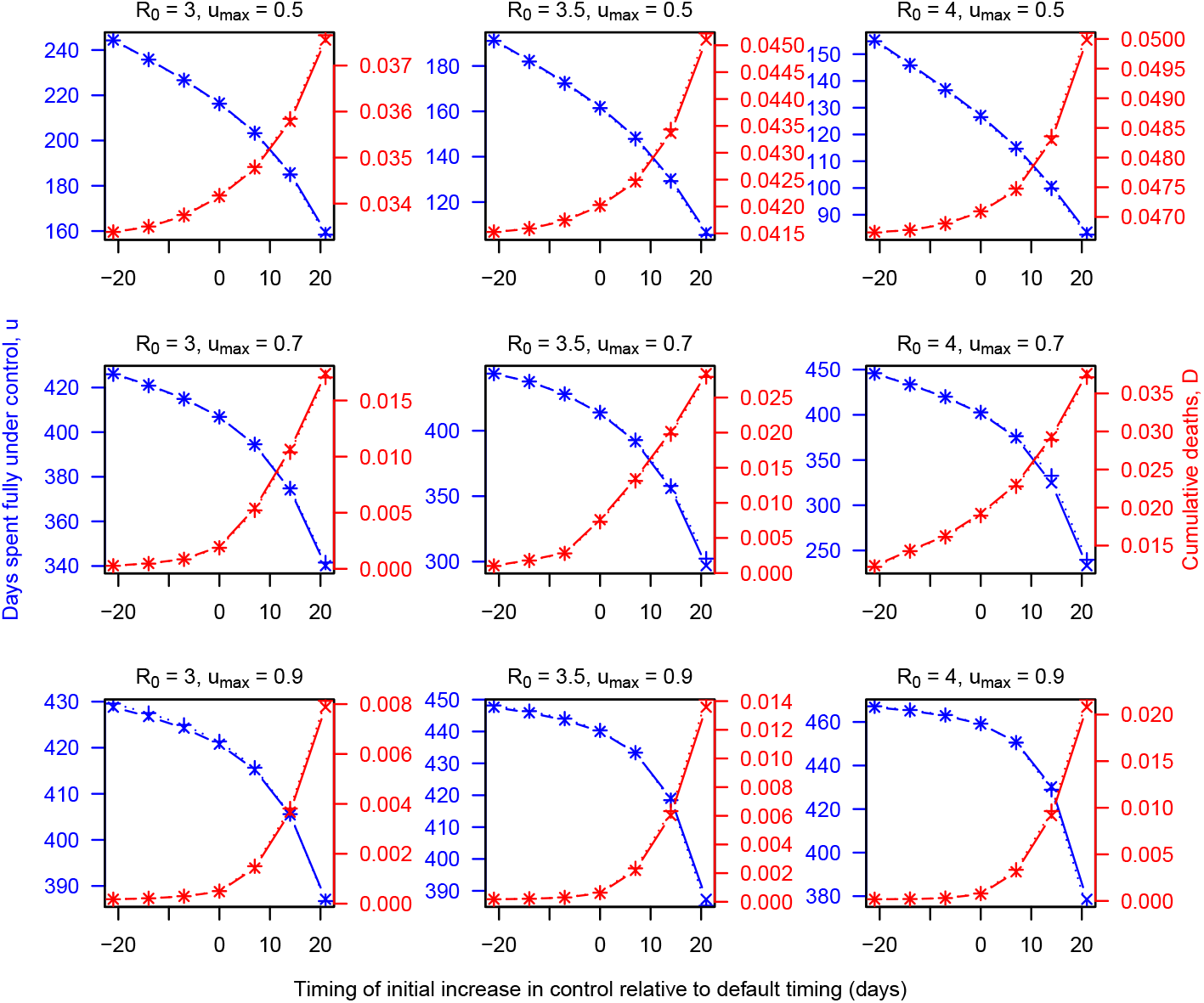
Dependence of time under control (blue) and cumulative deaths (red) on a shift in the timing of *u*(*t*) before April 30, 2020 (x-axis), *R*_0_ (columns), *u*_max_ (rows), and *ω* (markers). Deaths are quantified as a proportion of the overall population.

The overall amount of time spent under control throughout 2020 and 2021 increases when control begins earlier and decreases when control begins later (Figure 10, blue). This is the case across all combinations of *R*_0_, *u*_max_, and *ω* that we considered. In part, this owes to less time spent under control through April 30, 2020, when *u*(*t*) is fixed and not subject to optimization. In addition though, delays in the initiation of control result in a higher prevalence of infection by the beginning of the optimization period, which results in higher levels of subsequent transmission, greater depletion of the susceptible population, and less need for control later in the period of optimization (compare Figure 11 with Figure 5).

**Fig. 11.**
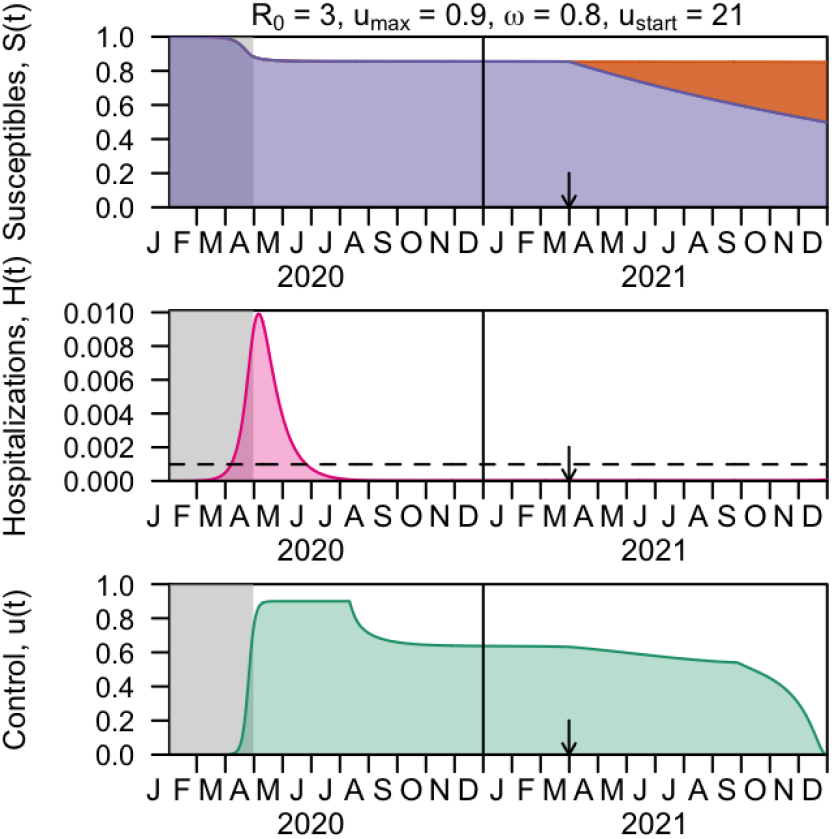
Optimal control (bottom) and its impacts on the dynamics of hospitalized (middle) and susceptible (top) compartments with *R*_0_ = 3, *u*_max_ = 0.9, *ω* = 0.8, *c* = 10^−12^, and the initiation of control delayed by 21 days. Vaccination is introduced at the time indicated by the arrow, with the vaccinated population (orange) reducing the susceptible population. Through April 2020 (gray shading), the value of the control is fixed according to its calibrated trajectory. The dashed horizontal line indicates hospital capacity.

## 4 Discussion

Under our model, the ability of NPIs to minimize deaths depends to a large extent on the maximum effect that NPIs could have on transmission, as captured by the parameter *u*_max_. If *u*_max_ is high (0.9), our model predicts that maintaining transmission at a low enough level that hospital capacity will not be exceeded could be possible. If *u*_max_ is low (0.5), our model predicts that there may be limited scope to curtail the pandemic, even if control is sustained for a long period of time. If *u*_max_ is intermediate (0.7), our model predicts that the potential impact of control is sensitive to the value of *R*_0_. These results emphasize the importance of careful estimation of these parameters as the pandemic progresses [18]. This includes accounting for geographic differences in both *u*_max_ and *R*_0_ [19,21].

The balance between minimizing deaths versus days under control is determined by the parameter *c* in our model, which mirrors the weighting between these factors that government leaders will need to consider as they make decisions in coming months. In the event that NPIs are effective, our analysis shows that they will need to be sustained at a high level until sometime in summer 2020, at which time they can be relaxed to a small degree but will still need to be maintained at a relatively high level thereafter. Scenarios that place greater weight on minimizing days under control provide insight into the possible consequences of relaxing NPIs prematurely. In a scenario in which the effect of NPIs is reduced moderately in May, resuming NPIs at high levels soon after becomes necessary to react to a second wave later in the summer. In a scenario in which NPIs are reduced more drastically in May, an extremely large second wave occurs in summer 2020 that exceeds hospital capacity many times over.

Our conclusion that prolonged control is needed to avoid a resurgence that would greatly exceed healthcare capacity is in line with results from other modeling studies [13,37,17,25,51]. Some differ though in that they consider scenarios in which control is implemented intermittently, with periods of relaxed control in between periods of maximal control [17,25,51]. In the limited number of direct comparisons of intermittent and continuous strategies that have been performed for COVID-19 to date, continuous strategies appear to be capable of more effectively limiting transmission to low levels [14,52]. The sustainability of either strategy would benefit from the ability to transition away from heightened social distancing and more toward diagnostic testing, contact tracing, and case isolation [51,43].

Our results tend to agree with those from other optimal control analyses of COVID-19, although our analysis goes beyond those studies in some ways. In general, ours and other studies share the conclusion that heightened control early in the pandemic is important for achieving long-term success. Like our analysis, some anticipate that partial relaxation of controls may be possible over time [14,45], whereas others focus on strategies intended to have a more limited duration to begin with [40,35,43]. Importantly though, our analysis goes further than others in exploring sensitivity of the optimal control to model parameters. Specifically, we show that preventing a large wave that overwhelms health systems may not even be possible under some parameter combinations (low *u*_max_, high *R*_0_) and that prioritizing the minimization of deaths versus days under control leads to vastly different outcomes. We also constrain levels of control applied through April 2020, making the optimization more relevant to future decision making. Shifting the timing of the initiation of control shows that constraints about past levels of control strongly affect future possibilities for the extent to which deaths can be minimized and the level of control required to achieve that, consistent with other results [27].

The goal of our analysis is to provide qualitative insights into the implications of alternative approaches to control, rather than to make quantitative predictions about future events. Some key limitations of using our model for the latter purpose include our omission of subnational variation in epidemic dynamics [42,49], differentiation among alternative NPIs [18], and age differences in contact patterns [23] and risk of hospitalization [9]. At least with respect to the possibility of different levels of control for different age groups, one study found that age-specific optimal controls were all relatively similar for NPIs against pandemic influenza [46]. Additional limitations that affect our model’s suitability for making future predictions include its deterministic nature and the rudimentary calibration procedure that we performed, which was sufficient to provide a basis for qualitative analyses but that would need refinement for application of our model to inference or forecasting.

In conclusion, our analysis suggests that we are at a critical juncture in the pandemic, when decisions about the continuation or relaxation of NPI-based control strategies could have major implications for the possibility of keeping transmission below levels that health systems can cope with. At the same time, our analysis highlights the role that constraints play in determining optimal levels of control going forward, both in terms of constraints on epidemiological parameters and on levels of control prior to the time that a decision is made about future actions. Going forward, reducing transmission in the near term would give decision makers greater flexibility in the range of decisions available to them in the future, and gathering high-quality data could help reduce uncertainty about the consequences of those decisions.

## Data Availability

All code and data used in our analysis is available at http://github.com/TAlexPerkins/covid19optimalcontrol.

## Acknowledgements

Thanks to the University of Notre Dame’s Center for Research Computing for computing resources.

## Notes

The authors were supported by a RAPID grant from the National Science Foundation (DEB 2027718).

### Competing Interest Statement

The authors have declared no competing interest.

### Funding Statement

The authors were supported by a RAPID grant from the National Science Foundation (DEB 2027718).

## References

1. nCoV 2019 Data Working Group: Epidemiological Data from the nCoV-2019 Outbreak: Early Descriptions from Publicly Available Data (2020 (accessed April 15, 2020)). URL http://virological.org/t/epidemiological-data-from-the-ncov-2019-outbreak-early-descriptions-from-publicly-available-data/337

2. Agusto, F., Khan, M.: Optimal control strategies for dengue transmission in pakistan. Mathematical Biosciences 305, 102–121 (2018). DOI https://doi.org/10.1016/j.mbs.2018.09.007. URL http://www.sciencedirect.com/science/article/pii/S002555641830453X

3. Aledort, J., Lurie, N., Wasserman, J., Bozzette, S.: Non-pharmaceutical public health interventions for pandemic influenza: an evaluation of the evidence base. BMC Public Health 7, 208 (2007)

4. Anderson Cooper 360, CNN: Lead scientist says coronavirus vaccine could be ready soon (2020 (accessed April 15, 2020)). URL https://www.cnn.com/videos/health/2020/04/15/coronavirus-vaccine-timeline-acfc-vpx.cnn

5. Bedford, J., Enria, D., Giesecke, J., Heymann, D., Ihekweazu, C., Kobinger, G., Lane, H., Memish, Z., Oh, M., Sall, A., Schuchat, A., Ungchusak, K., Wieler, L.: Covid-19: towards controlling of a pandemic. Lancet (2020). DOI 10.1016/S0140-6736(20)30673-5

6. Blayneh, K., Gumel, A., Lenhart, S., Clayton, T.: Backward bifurcation and optimal control in transmission dynamics of west nile virus. Bulletin of Mathematical Biology 72, 1006–1028 (2010)

7. Centers for Disease Control and Prevention: 2009 H1N1 Flu Vaccine (2010 (accessed April 15, 2020)). URL https://www.cdc.gov/h1n1flu/vaccination/

8. Centers for Disease Control and Prevention: 2009 H1N1 Pandemic Timeline (2010 (accessed April 15, 2020)). URL https://www.cdc.gov/flu/pandemic-resources/2009-pandemic-timeline.html

9. Centers for Disese Control and Prevention: Severe outcomes among patients with corossnavirus disease 2019 (covid-19) - united states, february 12-march 16 2020. MMWR Morb Mortal Wkly Rep 2020 69, 343–346 (2020). DOI http://dx.doi.org/10.15585/mmwr.mm6912e2

10. Chinazzi, M., Davis, J.T., Ajelli, M., Gioannini, C., Litvinova, M., Merler, S., Pastore y Piontti, A., Mu, K., Rossi, L., Sun, K., Viboud, C., Xiong, X., Yu, H., Halloran, M.E., Longini, I.M., Vespignani, A.: The effect of travel restrictions on the spread of the 2019 novel coronavirus (covid-19) outbreak. Science (2020). DOI 10.1126/science.aba9757. URL https://science.sciencemag.org/content/early/2020/03/05/science.aba9757

11. Choi, S., Jung, E.: Optimal tuberculosis prevention and control strategy from a mathematical model based on real data. Bulletin of Mathematical Biology 76, 1566–1589 (2014)

12. Cowling, B., Ali, S., Ng, T., Tsang, T., Li, J., Fong, M., Liao, Q., Kwan, M., Lee, S., Chiu, S., Wu, J., Wu, P., Leung, G.: Impact assessment of non-pharmaceutical interventions against coronavirus disease 2019 and influenza in hong kong: an observational study. Lancet Public Health (2020). DOI 10.1016/S2468-2667(20)30090-6

13. Davies, N., Kucharski, A., Eggo, R., Gimma, A., Edmunds, W.: The effect of non-pharmaceutical interventions on COVID-19 cases, deaths and demand for hospital services in the UK: a modeling study (2020). URL https://cmmid.github.io/topics/covid19/control-measures/uk-scenario-modelling.html

14. Djidjou-Demasse, R., Michalakis, Y., Choisy, M., Sofonea, M., Alizon, S.: Optimal covid-19 epidemic control until vaccine deployment. medRxiv (2020). DOI 10.1101/2020.04.02.20049189

15. van den Driessche, P., Watmough, J.: Further notes on the basic reproduction number. In: F. Brauer, P. van den Driessche, J. Wu (eds.) Mathematical Epidemiology, pp. 159–178. Springer (2008)

16. Editorial: Covid-19 in the usa: a question of time. Lancet 395, 1229 (2020)

17. Ferguson, N., Laydon, D., Nedjati-Gilani, G., Imai, N., Ainslie, K., et al.: Report 9 - Impact of non-pharmaceutical interventions (NPIs) to reduce COVID-19 mortality and healthcare demand (2020). URL https://www.imperial.ac.uk/mrc-global-infectious-disease-analysis/covid-19/report-9-impact-of-npis-on-covid-19/

18. Flaxman, S., Mishra, S., Gandy, A., Hjt, U., Coupland, H., Mellan, T., et al.: Report 13 - Estimating the number of infections and the impact of nonpharmaceutical interventions on COVID-19 in 11 European countries (2020). URL https://www.imperial.ac.uk/mrc-global-infectious-disease-analysis/covid-19/report-13-europe-npi-impact/

19. Gilbert, M., Pullano, G., Pinotti, F., Valdano, E., Poletto, C., Boelle, P., D’Ortenzio, E., Yazdanpanah, Y., Eholie, S., Altmann, M., Gutierrez, B., Kraemer, M., Colizza, V.: Preparedness and vulnerability of african countries against importations of covid-19: a modelling study. Lancet 395, 871–877 (2020)

20. Google: COVID-19 Community Mobility Report (2020 (accessed April 15, 2020)). URL https://www.gstatic.com/covid19/mobility/2020-04-11_US_Mobility_Report_en.pdf

21. Hilton, J., Keeling, M.: Estimation of country-level basic reproductive ratios for novel coronavirus (covid-19) using synthetic contact matrices. medRxiv (2020). DOI 10.1101/2020.02.26.20028167

22. Holshue, M.L., DeBolt, C., Lindquist, S., Lofy, K.H., Wiesman, J., Bruce, H., Spitters, C., Ericson, K., Wilkerson, S., Tural, A., Diaz, G., Cohn, A., Fox, L., Patel, A., Gerber, S.I., Kim, L., Tong, S., Lu, X., Lindstrom, S., Pallansch, M.A., Weldon, W.C., Biggs, H.M., Uyeki, T.M., Pillai, S.K.: First case of 2019 novel coronavirus in the united states. New England Journal of Medicine 382(10), 929–936 (2020). DOI 10.1056/NEJMoa2001191

23. Jarvis, C., van Zandvoort, K., Gimma, A., Prem, K., Klepac, P., Rubin, G., Edmunds, W.: Quantifying the impact of physical distance measures on the transmission of covid-19 in the uk. medRxiv (2020). DOI 10.1101/2020.03.31.20049023

24. Keeling, M., Rohani, P.: Modeling Infectious Diseases in Humans and Animals. Princeton University Press (2007)

25. Kissler, S., Tedijanto, C., Goldstein, E., Grad, Y., Lipsitch, M.: Projecting the transmission dynamics of sars-cov-2 through the postpandemic period. Science p. eabb5793 (2020)

26. Kraemer, M., Yang, C., Gutierrez, B., Wu, C., Klein, B., Pigott, D., et al.: The effect of human mobility and control measures on the covid-19 epidemic in china. Science p. eabb4218 (2020)

27. Lai, S., Ruktanonchai, N., Zhou, L., Prosper, O., Luo, W., Floyd, J., Wesolowski, A., Santillana, M., Zhang, C., Du, X., Yu, H., Tatem, A.: Effect of non-pharmaceutical interventions for containing the covid-19 outbreak in china. medRxiv (2020). DOI 10.1101/2020.03.03.20029843

28. Lenhart, S., Workman, J.T.: Optimal Control Applied to Biological Models. Chapman and Hall/CRC (2007)

29. Lin, F., Muthuraman, K., Lawley, M.: An optimal control theory approach to non-pharmaceutical interventions. BMC Infectious Diseases 10, 32 (2010)

30. Lurie, N., Saville, M., Hatchett, R., Halton, J.: Developing covid-19 vaccines at pandemic speed. New England Journal of Medicine (2020).DOI 10.1056/NEJMp2005630

31. Mallela, A.: Optimal control applied to a seir model of 2019-ncov with social distancing. medRxiv (2020). DOI 10.1101/2020.04.10.20061069

32. Martin, J., Hamilton, B., Osterman, M., Driscoll, A.: Births: Final data for 2018. National Vital Statistics Reports 68, 13 (2019)

33. Miller Neilan, R., Scaefer, E., Gaff, H., Fister, H., Lenhart, S.: Modeling optimal intervention strategies for cholera. Bulletin of Mathematical Biology 72, 2004–2018 (2010)

34. Mizumoto, K., Kagaya, K., Zarebski, A., Chowell, G.: Estimating the asymptomatic proportion of coronavirus disease 2019 (covid-19) cases on board the diamond princess cruise ship, yokohama, japan, 2020. Eurosurveillance 25, 2000180 (2020)

35. Morris, D., Rossine, F., Plotkin, J., Levin, S.: Optimal, near-optimal, and robust epidemic control. arXiv (2020). DOI arXiv:2004.02209

36. New York Times: Coronavirus (Covid-19) Data in the United States (2020 (accessed April 15, 2020)). URL https://github.com/nytimes/covid-19-data

37. Ngoonghala, C., Iboi, E., Eikenberry, S., Scotch, M., MacIntyre, C., Bonds, M., Gumel, A.: Mathematical assessment of the impact of non-pharmaceutical interventions on curtailing the 2019 novel coronavirus. medRxiv (2020). DOI 10.1101/2020.04.15.20066480

38. Onder, G., Rezza, G., Brusaferro, S.: Case-Fatality Rate and Characteristics of Patients Dying in Relation to COVID-19 in Italy. JAMA (2020). DOI 10.1001/jama.2020.4683. URL https://doi.org/10.1001/jama.2020.4683

39. Park, S., Bolker, B., Champredon, D., Earn, D., Li, M., Weitz, J., Grenfell, B., Dushoff, J.: Reconciling early-outbreak estimates of the basic reproductive number and its uncertainty: framework and applications to the novel coronavirus (sars-cov-2) outbreak. medRxiv (2020). DOI 10.1101/2020.01.30.20019877

40. Patterson-Lomba, O.: Optimal timing for social distancing during an epidemic. medRxiv (2020). DOI 10.1101/2020.03.30.20048132

41. Perkins, T., Cavany, S., Moore, S., Oidtman, R., Lerch, A., Poterek, M.: Estimating unobserved sars-cov-2 infections in the united states. medRxiv (2020). DOI 10.1101/2020.03.15.20036582

42. Perkins, T., Rodriguez-Barraquer, I., Manore, C., Siraj, A., Espana, G., Barker, C., Johansson, M., Reiner, R.: Heterogeneous local dynamics revealed by classification analysis of spatially disaggregated time series data. Epidemics p. 100357 (2019)

43. Piguillem, F., Shi, L.: Optimal covid-19 quarantine and testing policies. EIEF Working Papers Series 2004, Einaudi Institute for Economics and Finance (2020). URL https://ideas.repec.org/p/eie/wpaper/2004.html

44. Prem, K., Liu, Y., Russell, T., Kucharski, A., Eggo, R., Davies, N., Jit, M., Klepac, P.: The effect of control strategies to reduce social mixing on outcomes of the covid-19 epidemic in wuhan, china: a modelling study. Lancet Public Health (2020). DOI 10.1016/S2468-2667(20)30073-6

45. Shah, N., Suthar, A., Jayswal, E.: Control strategies to curtail transmission of covid-19. medRxiv (2020). DOI 10.1101/2020.04.04.20053173

46. Shim, E.: Optimal strategies of social distancing and vaccination against seasonal influenza. Mathematical Biosciences and Engineering 10, 1615–1634 (2013)

47. Soetaert, K., Petzoldt, T., Setzer, R.W.: Solving differential equations in r: Package desolve. Journal of Statistical Software 33(9), 1–25 (2010). DOI 10.18637/jss.v033.i09. URL http://www.jstatsoft.org/v33/i09

48. Tchuenche, J., Khamis, S., Agustto, F., Mpeshe, S.: Optimal control and sensitivity analysis of an influenza model with treatment and vaccination. Acta Biotheoretica 59, 1–28 (2011)

49. Team, C.C..R.: Geographic differences in covid-19 cases, deaths, and incidence — united states, february 12–april 7, 2020. MMWR Morb Mortal Wkly Rep 69, 465–471 (2020)

50. health service utilization forecasting team, I.C.., Murray, C.J.: Forecasting covid-19 impact on hospital bed-days, icu-days, ventilator-days and deaths by us state in the next 4 months. medRxiv (2020). DOI 10.1101/2020.03.27.20043752. URL https://www.medrxiv.org/content/early/2020/03/30/2020.03.27.20043752

51. Tuite, A., Fisman, D., Greer, A.: Mathematical modelling of covid-19 transmission and mitigation strategies in the population of ontario, canada. Canadian Medical Association Journal 192, cmaj.200476 (2020)

52. Yap, W., Raja, D.: Time-variant strategies for optimizing the performance of non-pharmaceutical interventions (npis) in protecting lives and livelihoods during the covid-19 pandemic. medRxiv (2020). DOI 10.1101/2020.04.13.20063248

